# Estimation of reproduction number of SARS-CoV-2 Omicron variant outbreak in Hong Kong

**DOI:** 10.1101/2023.01.21.23284868

**Authors:** Mattia Allieta, Jelena Komloš, Davide Rossi Sebastiano

**Author notes:** **Corresponding author** Davide Rossi Sebastiano, MD, PhD, Neurophysiology Unit, Fondazione IRCCS Istituto Neurologico Carlo Besta, 20133 Milan, Italy. Mail. **Author contributions Mattia Allieta:** conceptualization (lead); data curation (equal); formal analysis (lead); writing-original draft (supporting); writing-review and editing (supporting). **Jelena Komloš:** data curation (equal); writing-review and editing (lead). **Davide Rossi Sebastiano:** writing-original draft (lead); supervision (lead). **Funding:** This research received no external funding. **Patients Involvement:** No patients were involved.

## Abstract

We present the evolution of time dependent reproduction number across the five different Hong Kong Special Administrative Region of the People’s Republic of China (HK) epidemic waves from January, 2020, to March, 2022. We provide reliable estimation of reproduction number of Omicron variant of concern (VOC) by analysing data related to fifth wave to determine its peculiar characteristics with respect to the other VOCs.

HK could be considered as the optimal model for the calculation of the dynamics of Omicron VOC transmission in an environment representative of the fully populated cities of the Asian Pacific coast.

On the basis of Rt calculated for Omicron VOC in our work, researchers could refine provisional data for the current outbreak which is affecting China

## 1. Introduction

The Hong Kong Special Administrative Region of the People’s Republic of China (HK) recorded relatively few cases of coronavirus disease 2019 (COVID-19), and especially if compared with other densely populated regions, due to the quick restrictive measures adopted at the beginning of the outbreak, which caused low local transmission rates with few or no local infections, as demonstrated by the mean effective reproductive number between January 23^rd^, 2020 and May 12^th^, 2021 **[1,2]**. As a result, from January 2020, HK had a total cumulative incidence of about 10000 positive tested cases thanks to an “ elimination strategy” which supports the so called “ Zero-COVID” regime, albeit it featured formally four different pandemic waves.

However, in January 2022 the sudden increase of new positive tested cases boosted the fifth wave of epidemic, with such a worrying increase in new cases that it has pushed China’s President Xi Jinping to order HK to stabilize its covid-19 fifth wave of epidemic as “ overriding mission” [3]. The new wave was attributed to the B.1.1.529 variant of severe acute respiratory syndrome coronavirus 2 (SARS-CoV-2), designated as variant of concern (VOC) Omicron by the World Health Organization (WHO) on November 26^th^, 2021 **[4]**.

Since HK Chief Executive Carrie Lam decided to track the epidemic by an extensive and enduring contact tracing involving the entire HK population, the fifth HK wave offered a unique way to determine quantitative epidemiological parameter related to the Omicron VOC only, because the most of the recorded infections have been attributed only to Omicron VOC which developed from an environment where a “ Zero COVID” regime was present and other VOCs were not existing. Since China’s recent COVID-19 outbreak is predominantly led by the Omicron subvariants BA.5.2 and BF.7, which together account for 97.5% of all local infections **[5]**, the analysis of HK dynamics of 2022 epidemic could represent an effective model for it.

In a previous work, we provided a mapping of the Alpha VOC transmission dynamics spread in all regions of Italy in the first year of COVID-19 pandemic **[6]**. With the same methods, we studied the evolution of time dependent reproduction number across the five different HK epidemic waves to obtain an estimation of reproduction number of Omicron VOC in order to determine its peculiar characteristics with respect to the other VOCs.

## 2. Material and Methods

### 2.1. Epidemiological data

Official data of COVID-19 pandemic for Hong-Kong has been taken from the complete *<u>Our World in Data</u>* COVID-19 dataset as downloaded by https://ourworldindata.org/ website. These data are available as open source for all purposes and complemented by vaccination data as implemented by Mathieu et a., 2021 **[7]**. Data for the analysis were considered from 2020-01-23 to 2022-03-01, i.e. 769 days from the onset related to the first COVID-19 positive cases recorded in Hong-Kong. In this period, we consider the daily number of new confirmed positive cases defined as “ Daily Incidence” and the number of Fully Vaccinated people, i.e. the number of people that received a full vaccination cycle.

### 2.2 Estimation of time dependent reproduction number *R*_*t*_

To evaluate the time dependent reproduction number *R*_*t*_ we adopted the method developed by Wallinga and Teunis, 2004 **[8]**.

The transmission probability (*p*_*ij*_) of individual *i* being infected by individual *j* at *t*_*i*_, *t*_*j*_ onsets, respectively, can be described mathematically as **[9]**:

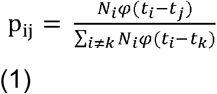

where *φ*_*i*_ is the distribution of the generation time corresponding to the distribution of the serial interval, i.e. the time between when a person gets infected and when they subsequently infect another other people, calculated at time *i* within the assumption that the incubation period does not change over the course of the epidemic **[10]**. We considered that the distribution of the serial interval was expected to follow a gamma distribution with mean (±SD) of 6.50 ± 4.03 days as reported by the Imperial College COVID-19 Response Team **[11]**.

The net reproduction number *R*_j_ is then then sum of all *p*_*ij*_ involving *j* as the infector *R*_*j*_ =*∑*_*j*_ *p*_*ij*_ and it can be averaged over all cases with same date of onset as 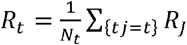.

Since *R*_*t*_ are computed by averaging over all transmission networks compatible with observed incidence data, no assumption is made about the time dependence of the epidemic unlike, for example the exponential growth in the well-known Bayesian approach **[8,9]**.

We believe, hence, that this model is particularly suitable to estimate the reproduction number in the post-peak period where the transmission is expected to decrease. All the above data analyses were performed using the R_0_ package **[9]** as implemented in statistical software R (R Core Team; R: A language and environment for statistical computing. R Foundation for Statistical Computing, Vienna, Austria. URL https://www.R-project.org/], 2017).

## 3. Results

The daily incidence of COVID-19 tested positive cases in HK is represented in Figure 1.

**Figure 1.**
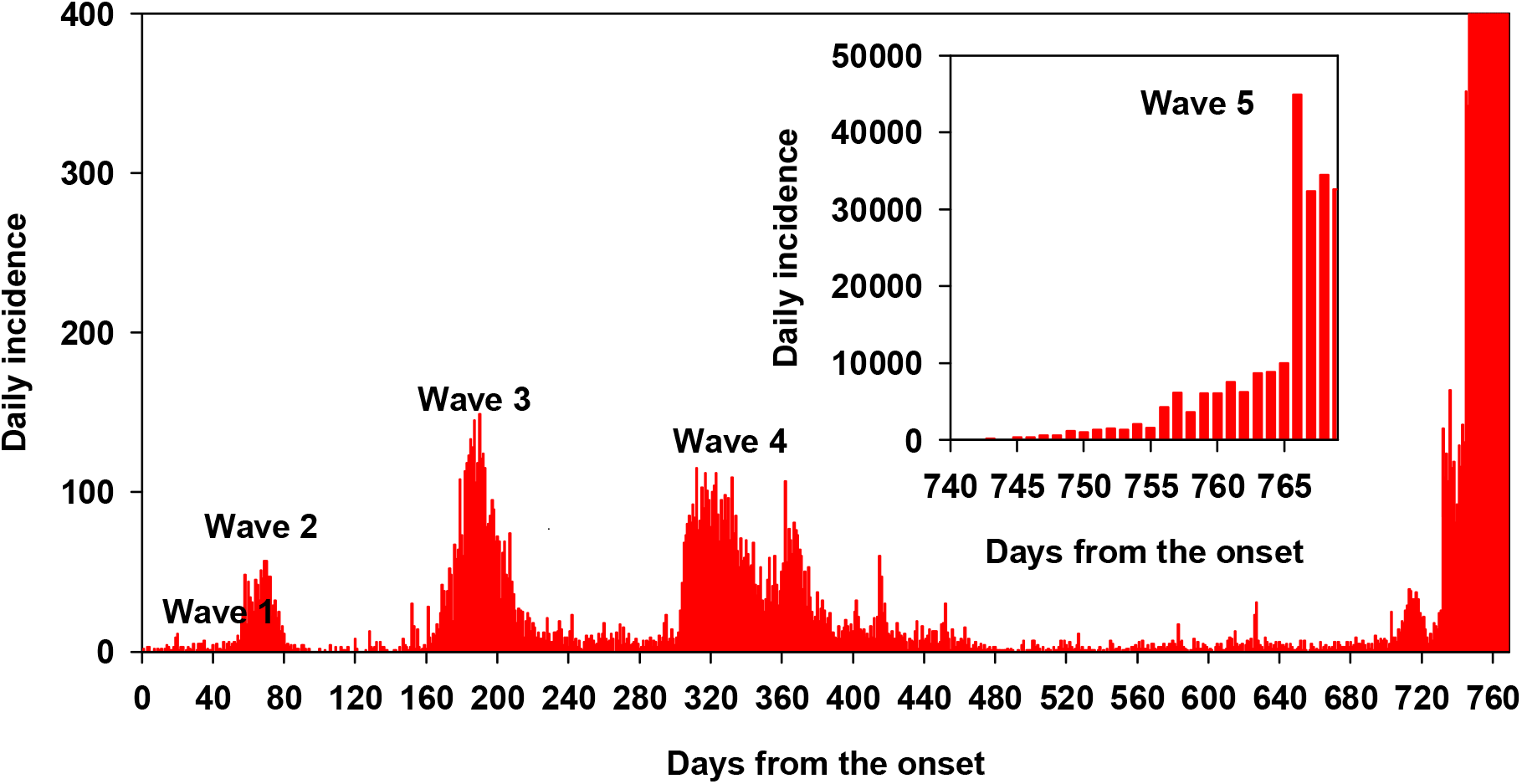
Daily incidence related to confirmed cases in Hong Kong.

From 2020, January 1^st^, to 2022, March 1^st^, HK featured different five pandemic waves. Looking at the smoothed time dependence of effective reproduction number across the waves (Figure 2), we recorded a rather modest variation with only few events weakly deviating from the control *R*_*t*_ = 1 regime. However, at the beginning of January, 2022 the sudden increase of daily incidence boosted dramatically the fifth wave of epidemic reaching the 50,000 cases in few days, with maximal Rt and smoothed Rt estimated reaching about 3.5 and 2.2, respectively (see also Figure 2)

**Figure 2.**
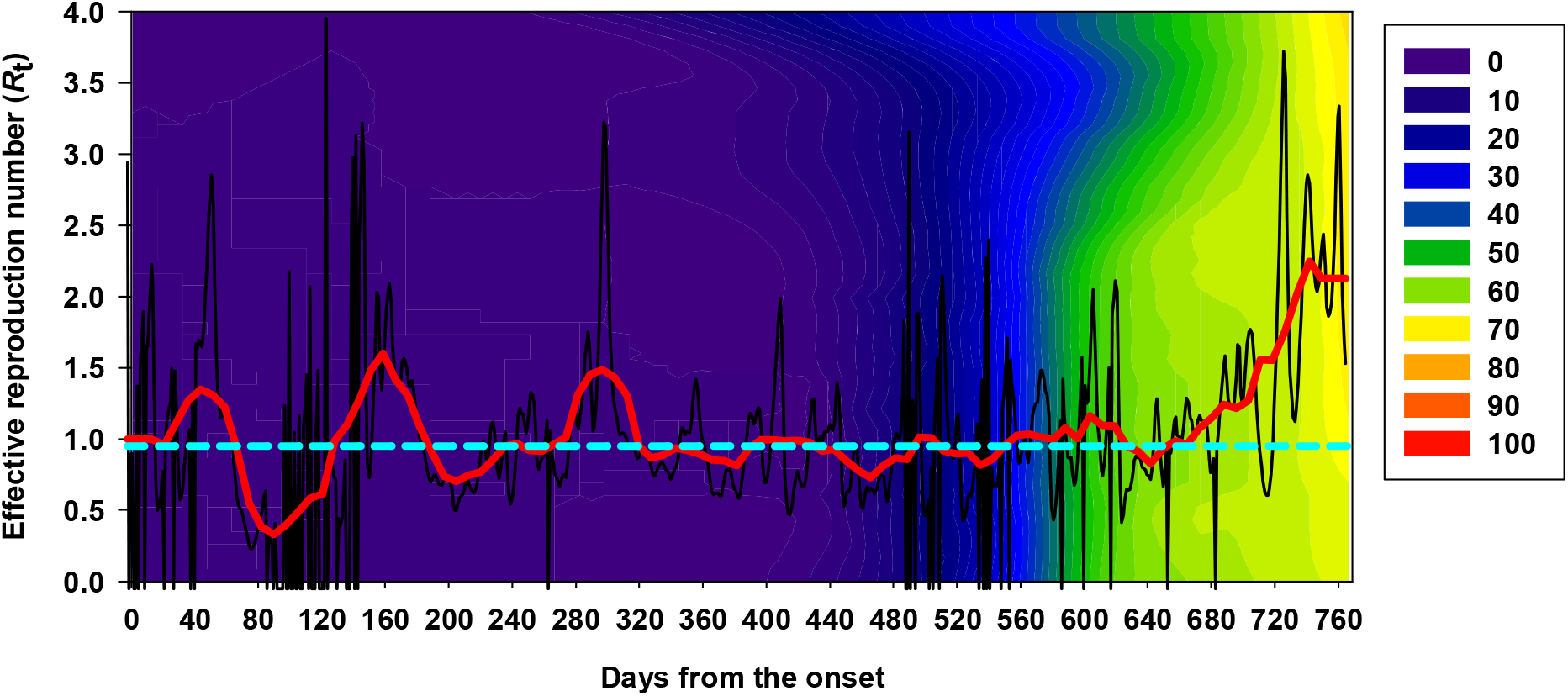
Time evolution of effective reproduction number across the five waves: original series (thinner profile) and moving average smoothing (thicker profile). The false color scale represents the percentage (%) of fully vaccinated people over the total population resident in Hong Kong **[7]**.

## 4. Discussion

Based on previous literature, we know that Omicron VOC showed early doubling time consistently shorter than Beta and Delta VOC **[12]**, with a reproduction number expected greater more than three times with respect to Delta **[13]**. However, Omicron VOC epidemic developed in countries where other VOCs are highly diffused, so it is difficult to derive the contribution of Omicron VOC only from the analysis of cumulative positive cases recorded without an accurate “ genomic” tracing able to separate the relative contribution of several VOCs. Hence, only few estimations are available in the literature, based mainly by defining the ratio with respect to Delta VOC or by analysing small cluster of infected people **[14-18]**.

Despite HK is one of the most densely populated area in the world, before 2022, the effects of restrictive measures imposed by HK government were very satisfactory in limiting the spread of pandemic, despite the virus mutations **[19**,**20]**, hence the fifth wave in HK offered a different perspective to evaluate intrinsic epidemiological parameters related to Omicron VOC because it developed from a rather unperturbed zero-covid environment.

In this work we estimated the reproduction number of Omicron VOC by means of the analysis of the fifth wave of COVID-19 pandemic in HK, featured since January to February, 2022, determining and comparing the evolution of time dependent reproduction number across the five different HK waves in order to obtain the peculiar characteristics of Omicron VOC with respect to the other epidemic waves. We determined for the fifth HK pandemic (highly related to Omicron VOC) a mean Rt approx. 2, in agreement with Kim et al. **[21]**.

Omicron VOC has a growth advantage over the others because of its higher transmissibility, immune evasion and shorter serial interval **[15]**, which is shorter than or close to its median incubation period **[22]**. This suggests that a significant amount of secondary transmission may occur prior to the symptomatic disease onset, thus facilitating the epidemic. In the fifth wave of HK, the effective Rt remained persistently high even in a context with relevant fully vaccination coverage **[23]** (see also the “ false colours” contour plot in Figure 1), probably due to the scarce effectiveness of BNT162b2 and CoronaVac against Omicron VOC, especially in children and adolescents **[24]**.

## 5. Conclusions

The initial stages of the fifth wave of the COVID-19 pandemic in HK, which occurred in January 2022, could be considered as the optimal model for the calculation of the dynamics of Omicron VOC transmission in an environment representative of the fully populated cities of the Asian Pacific coast.

Moreover, on the basis of Rt calculated for Omicron VOC in our work, researchers could refine provisional data for the current outbreak which is affecting China.

## Data Availability

All data produced in the present work are contained in the manuscript

## Notes

**Conflicts of Interest:** Authors declare no conflicts of interest

### Competing Interest Statement

The authors have declared no competing interest.

### Funding Statement

This study did not receive any funding

### Author Declarations

The study used ONLY openly available human data that were originally located at Our World in Data https://ourworldindata.org/

